# Electrocardiographic Changes After Completion of a Triathlon

**DOI:** 10.1101/19000687

**Authors:** Caroline Hosatte-Ducassy, José A. Correa, François Lalonde, Rohit Mohindra, Gregory Marton, Michael Chetrit, Audrey Marcotte, François Tournoux, Eileen Bridges

## Abstract

**Objectives:** Given the increasing popularity of long-distance triathlon events amongst amateur athlete and the difficulty for emergency physician to address cardiovascular complaints in the context of exercise, this study aims to:

- Identify the prevalence of electrocardiographic abnormalities before and after a long distance triathlon in a cohort of participants using the Seattle criteria.
- Identify the acute changes that occur on their ECGs at the finish line of a long-distance triathlon.

**Methods:** This prospective observational study examines the prevalence of selected standard 12-lead ECG findings, the Seattle criteria, in asymptomatic athletes before and after the completion of a long-distance triathlon.

**Results:** Of 99 ECGs obtained prior to the race, 28 were abnormal, for a pre-race prevalence of 28.3% (95% CI (20.4, 37.8)). Of the 72 ECGs post-race, 12 were abnormal, for a post-race prevalence of 16.7% (95% CI (9.8, 26.9)). We did not observe any athletes with marked repolarization abnormalities.

Common findings were increased QRS voltage significant for left ventricular hypertrophy (LVH) (24 (24.2%) pre-race, 10 (14.1%) post-race), early repolarization (21 (21.2%) pre-race, 19 (26.8%) post-race) and incomplete right bundle branch block (RBBB) (8 (8.1%) pre-race, 11 (15.5%) post-race).

McNemar’s test showed no agreement between the ECG pre and post results (Chi-square =6.54, *p* = 0.01), suggesting a possible effect of the race on ECG findings. We observed a trend to normalization of athlete’s ECGs with acute exercise.

**Conclusion:** Long-distance endurance exercise might acutely affect the ECGs findings in asymptomatic athletes and abnormal ECG findings were common in our cohort of athletes. Physicians providing care to long-distance athletes should interpret ECGs in this population prudently.

**SUMMARY BOX:**

- The acute effect of exercises on athlete’s electrocardiograms has not been well studied.
- In our cohort of long-distance triathlon finishers, 28.3% of athletes had abnormal ECGs pre-race and 16.7% had abnormal ECGs post-race according to the Seattle Criteria. We observed a trend toward normalization of athlete’s ECGs with acute exercise.
- Common ECGs abnormalities found in those asymptomatic athletes were left ventricular hypertrophy, early repolarization and right bundle branch block.
- Physicians involved in the care of athletes should be prudent when interpreting ECGs in this population.

## INTRODUCTION

The popularity of high-volume endurance athletic events surged in the last 30 years. Marathons, triathlons of all distances and obstacle courses attract an increasing number of participants every year.[1, 2] Amateur athletes are engaging in regular endurance training [2] and a proportion of these participants, with a mean age of 40 to 50 years old, have predisposing risk factors and may have a cardiac event during training or racing.[3]

The physiologic adaptation of the heart in athletes (“athlete’s heart”) [4] and risk factors for sudden cardiac death (SCD) during exercise are reported in the literature. The European Society of Cardiology (ESC) first established criteria for pre-participation screening electrocardiograms (ECG) in athletes.[5] To improve the specificity of these criteria and avoid false-positive results, the Seattle criteria were developed and are used for screening in athletes (see Table 1). [6-10] More recently, these criteria are refined to high-risk ECG findings, warranting more investigations, and low-risk findings, which, if found alone, most likely represent physiological response of the heart to exercise.[11] Attempts to correlate ECG changes to echocardiographic findings show limited predictive value of ECGs to reveal underlying cardiac abnormalities.[2, 12, 13]

**Table 1.** Description of Seattle Criteria [6].

|  |  |
| --- | --- |
| C1 | Sinus bradycardia |
| C2 | Sinus arrhythmia |
| C3 | Ectopic atrial rhythm |
| C4 | Junctional escape rhythm |
| C5 | 1° AV block |
| C6 | Mobitz I 2° AV block |
| C7 | Incomplete RBBB |
| C8 | Isolated QRS voltage for LVH (no non-voltage criteria or associated findings) |
| C9 | Early repolarisation |
| C10 | Concave ST-elevation |
| C11 | T-wave inversion |
| C12 | ST segment depression |
| C13 | Pathologic Q waves |
| C14 | Complete LBBB |
| C15 | Intraventricular conduction delay |
| C16 | Left axis deviation |
| C17 | Left atrial enlargement |
| C18 | Right ventricular hypertrophy pattern |
| C19 | Sinus bradycardia |
| C20 | Long QT interval |
| C21 | Short QT interval |
| C22 | Brugada-like ecg pattern |
| C23 | Profound sinus bradycardia |
| C24 | Atrial tachyarrhythmias |
| C25 | Premature ventricular contractions |
| C26 | Ventricular arrhythmias |

However, the focus has been on long-term changes of the athlete’s heart and there is a paucity of literature describing acute changes in athletes’ ECGs after endurance events. Evaluation of cardiac complaints in athletes is a challenge for physicians in the acute setting because interpretation of the ECG should be made in the context of exercise, but no specific criteria exist to differentiate benign normal variations secondary to exercise versus pathological changes. [3, 14, 15] Thus, athletes are prone to undergo unnecessary investigations [14] or might receive inappropriate reassurance or discharge when they present to an emergency setting with cardiac related complaints.

## OBJECTIVES

Given the increasing popularity of long-distance triathlon events amongst amateur athlete and the difficulty for emergency physicians to address cardiovascular complaints in the context of exercise, this study aims to:

## METHODOLOGY

This prospective observational study of a convenience sample of participants in a long-distance triathlon (1.8 km swim, 90 km bike and 21.1 km run) was designed to examine prevalence of selected ECG findings, commonly accepted as the standard to identify training-related ECG changes (the Seattle criteria) [6] and changes that may occur acutely following an endurance event. The study was approved by the Institutional Review Board of McGill. It was conducted June 26^th^ 2016 in Mont-Tremblant Quebec.

### Participants and participants involvement

A group of triathletes were consulted informally during the study design to assess the feasibility of the study and their willingness to participate in such an experiment. At race registration, held two and three days prior to the event, Canadian athletes older than 18 years were informed of the purpose of the study and asked to participate. Volunteers satisfying these inclusion criteria were enrolled and, after informed consent, given a study number and completed a questionnaire detailing demographic variables (c.f. Supplementary Material 1).

Participants with a history of acute coronary syndrome, pacemaker or defibrillator, use of medications that may affect ECG or echocardiographic findings, and women currently pregnant or who were pregnant in the last 12 weeks were excluded. Participants experiencing chest pain or syncope during or after the event were to be directed to the medical tent for evaluation and treatment and excluded from final analysis. [16, 17]

### Data collection

Physicians and paramedic students were trained to obtain 12-lead ECGs from participants in conformity with the American Heart Association (AHA) standards of quality.[18] ECGs were acquired on each participant at rest during study enrollment and again at the finish line with Philips HeartStart MRx monitor/defibrillator. Athletes were tracked electronically by bib number and, at the finish line, a member of the research team confirmed voluntary consent and bought participants to a designated area, close to the medical tent. The post-event ECG was performed within 15 minutes [2] of completion of the race in the same conditions as the pre-race ECGs. Technical quality and brief screening of ECGs for high-risk findings was done on-site by an emergency physician and EM resident (B and H-D). Participants were informed of their ECG results at the time of acquisition. Appropriate follow-up of concerning ECG findings was ensured by medical personnel on site (Supplementary Material 2). ECGs were stored in an electronic format (12-lead and rhythm strip) and identified by participant study number. Confidentiality of participant’s information was ensured by the research team.

### Data analysis

ECG results were stripped of all identification data in order for reviewers to be blinded to subjects’ demographics and acquisition time. Two emergency physicians and a cardiology fellow reviewed each ECG result independently and evaluated each Seattle criterion as either present or absent. For each criterion, the final evaluation retained was the one made by at least two of three reviewers.

Following interpretation, the ECGs were categorized as normal (≤1 Seattle Criteria C1-C10 present) or abnormal (2 or more Seattle Criteria C1-C10 present and/or any of the Seattle Criteria C11-C26 present). [7,8]

Demographic characteristics of participants are reported as counts and percentages for categorical variables and as mean and standard deviation (SD) for continuous variables.

The prevalence of abnormal ECG findings pre and post-race were estimated and reported with a 95% confidence interval (CI).[19]

Differences in demographic characteristics between those who presented changes in ECG findings between pre-race and post-race were investigated with Fisher’s exact test (FET) for categorical variables and *t*-test for continuous variables.

In order to investigate a possible effect of exercise on the presence of ECG findings, we performed a McNemar’s test to compare differences in proportions between normal and abnormal changes in ECG findings between pre-race and post-race evaluations.

## RESULTS

### Participants

103 adult volunteers meet the inclusion criteria and were recruited in the study. 99 had baseline ECGs and demographic data collected at race registration. Seven participants had concerning findings identified on their pre-race ECGs (Supplementary Material 3). Of these, none had significant past medical history of cardiovascular disease or were on any medications. Two had previous cardiac investigation including normal echocardiogram, one was referred to cardiology and had a normal echocardiogram prior to the race. The athletes were given explanations about the nature of the findings and risks of participation, and signed waivers to compete in the race. All participants chose to compete in the event, were asymptomatic and did not require medical assistance. Appropriate follow-up was arranged.

72 participants had ECGs done following completion of their race. Seven participants did not finish the race (c.f. Figure 1 - Flow chart), none for cardiac related reasons. 20 participants did not volunteer to present for the post-race ECG, none required medical attention. Demographic characteristics are detailed in table *2*.

**FIGURE 1.**
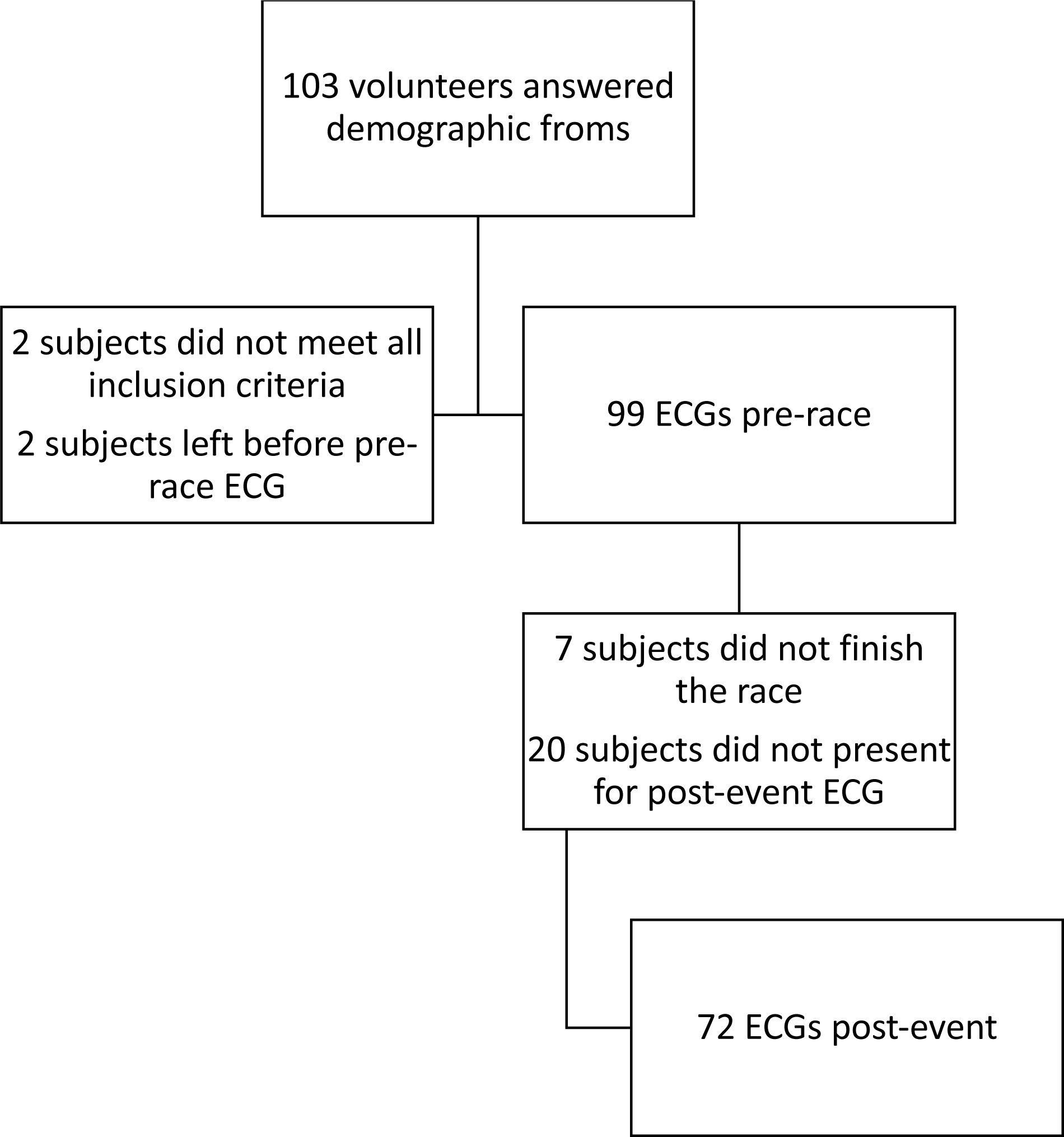
FLOW CHART OF PARTICIPANTS.

**Table 2.** Demographic characteristics of participants.

|  | *Pre-race (n = 99)* | *Post-race (n = 72)* |
| --- | --- | --- |
| *Age (years), mean (SD)* | *41.8 (10.2)* | *42.4 (10.2)* |
| *Sex (male), n (%)* | *58 (58.6%)* | *43 (59.7%)* |
| *Completion time (h:mm), mean (SD)* | *6h14 (1h54)* | *6h15 (1h07)* |
| *Number of hours training / week, n (%)* |  |  |
| *Less than 5 hours* | *6 (6.1%)* | *5 (6.9%)* |
| *5-15 hours* | *79 (79.8%)* | *57 (79.2%)* |
| *More than 15 hours* | *14 (14.1%)* | *10 (13.9%)* |
| *First long-distance athletic events (>4 hours), n (%)* | *25 (25.3%)* | *21 (29.2%)* |
| ***No participants had any significant past medical history or concerning symptoms during training.*** |  |  |

### Descriptive ECG findings

Of the 99 ECGs obtained pre-race, 28 (28.3%, 95% CI (20.4, 37.8)) were considered “abnormal” as defined by the Seattle criteria (with “normal” defined as no abnormalities or 1 low-risk abnormality identified). 36 (36.4%) had no Seattle criteria present. Seven (7.1%) ECGs were identified as “high-risk” by our reviewers. Details about those ECGs are available in supplementary material 3.

72 athletes presented for post-race ECGs. Of those, 48 (66.7%) had normal ECGs pre-race (≤ 1 Seattle Criteria C1-C10), including 25 (34.7%) with no Seattle criteria (C1-C26) present. Of the 24 (23.6%) abnormal, five (6.9%) had at least one high-risk finding present (C11-c26). The other abnormal pre-race ECGs were considered as such because of the presence of more than 1 low-risk feature, most commonly sinus bradycardia with benign early repolarization (n=4), left ventricular hypertrophy (n=5) or both (n=3).

Of the 72 ECGs post-race, 12 were abnormal (16.7%, 95% CI (9.8, 26.9)). In the 60 (83.3%) normal ECGs, 34 (47.2%) had no Seattle criteria (C1-C26) present. Five ECGs were interpreted as high risk, including two with persistence of pre-race findings, one with resolution of one finding and appearance of a long QT, and two participants had new ECG abnormalities. The prevalence for each individual Seattle Criteria is available in table 3.

**Table 3.** Prevalence of each Seattle criteria pre and post-race.

| Seattle Criteria |  | Numbers pre-race | Numbers of pre-race findings resolved post-race | Numbers post-race | Numbers of findings new at post-race |
| --- | --- | --- | --- | --- | --- |
| C1 | Sinus bradycardia | 26 | 24 | 2 | 0 |
| C2 | Sinus arrhythmia | 2 | 2 | 2 | 2 |
| C3 | Ectopic atrial rhythm | 0 | 0 | 0 | 0 |
| C4 | Junctional escape rhythm | 0 | 0 | 0 | 0 |
| C5 | 1° AV block | 0 | 0 | 0 | 0 |
| C6 | Mobitz I 2° AV block | 0 | 0 | 0 | 0 |
| C7 | Incomplete RBBB | 6 | 4 | 14 | 12 |
| C8 | Isolated QRS voltage for LVH (no non-voltage criteria or associated findings) | 17 | 10 | 11 | 4 |
| C9 | Early repolarisation | 17 | 7 | 20 | 10 |
| C10 | Concave ST-elevation | 0 | 0 | 0 | 0 |
| C11 | T-wave inversion | 1 | 1 | 0 | 0 |
| C12 | ST segment depression | 0 | 0 | 0 | 0 |
| C13 | Pathologic Q waves | 2 | 0 | 0 | 0 |
| C14 | Complete LBBB | 1 | 0 | 1 | 0 |
| C15 | Intraventricular conduction delay | 1 | 0 | 1 | 0 |
| C16 | Left axis deviation | 2 | 1 | 1 | 0 |
| C17 | Left atrial enlargement | 1 | 1 | 2 | 1 |
| C18 | Right ventricular hypertrophy pattern | 0 | 0 | 0 | 0 |
| C19 | Sinus bradycardia | 0 | 0 | 0 | 0 |
| C20 | Long QT interval | 0 | 0 | 3 | 3 |
| C21 | Short QT interval | 0 | 0 | 0 | 0 |
| C22 | Brugada-like ecg pattern | 0 | 0 | 0 | 0 |
| C23 | Profound sinus bradycardia | 0 | 0 | 0 | 0 |
| C24 | Atrial tachyarrhythmias | 0 | 0 | 0 | 0 |
| C25 | Premature ventricular contractions | 0 | 0 | 0 | 0 |
| C26 | Ventricular arrhythmias | 0 | 0 | 0 | 0 |

### ECG changes during the event

Table 4 shows a cross-classification of ECG results classified as Normal/Abnormal pre and post-race. There were 43 (59.7%) ECGs classified as Normal pre-race which remained Normal at post-race. ECG results did not coincide for 22 participants. As shown in table 4, 5 (6.9%) participants with normal ECG pre-race had abnormal ECG post-race ECGs and 17 (23.6%) participants with abnormal pre-race ECG had normal post-race ECG. McNemar’s test shows no agreement between the ECG pre and post results (Chi-square = 6.54, *p* = 0.01), suggesting a possible effect of the race on ECG findings.

**Table 4.** Cross-classification of pre and post-race ECG findings (n=72)

| Pre-race ECG | Post-race ECG |  |
| --- | --- | --- |
|  | Normal | Abnormal |
| Normal | 43 (59.7%) | 5 (6.9%) |
| Abnormal | 17 (23.6%) | 7 (9.7%) |

**Table 5.** Participant demographics with and without ECG findings pre to post-race (n=72)

|  | No change (n=50) | Change (n=22) | p value |
| --- | --- | --- | --- |
| Age (years), mean (SD) | 42.3 (9.9) | 42.6 (11.1) | 0.91* |
| Sex, n(%) |  |  |  |
| Male | 26 (52.0%) | 17 (77.3%) | 0.07** |
| Female | 24 (48.0%) | 5 (22.7%) |  |
| Training/week<sup>§</sup>, n(%) |  |  | 0.05** |
| < 5 hours | 5 (10.0%) | 0 |  |
| 5-15 hours | 41 (82.0%) | 16 (72.7%) |  |
| >15 hours | 4 (8.0%) | 6 (27.3%) |  |
| Number of years training for endurance event >4 hours, n(%) |  |  | 0.88** |
| <1 | 1 (2.0%) | 1 (4.6%) |  |
| 1-4 | 22 (44.0%) | 8 (36.4%) |  |
| 5-10 | 18 (36.0%) | 9 (40.9%) |  |
| >10 | 9 (18.0%) | 4 (18.2%) |  |
| Last time you exercised for more than 20 min. (Aerobic, moderate to intense), n(%) |  |  | 0.14** |
| >24 hours | 22 (44.0%) | 4 (18.2%) |  |
| In the last 24 hours | 13 (26.0%) | 9 (40.9%) |  |
| In the last 12 hours | 12 (24.0%) | 5 (22.7%) |  |
| < 2 hours ago | 3 (6.0%) | 4 (18.2%) |  |
| Prior participation in endurance athletic event lasting more than 4 hours, n(%) |  |  | 0.39** |
| No | 17 (34.0%) | 4 (18.2%) |  |
| In the last year | 25 (50.0%) | 13 (59.1%) |  |
| >1 year ago | 8 (16.0%) | 5 (22.7%) |  |
| Race time (min.), mean (SD) | 376.9 (48.7) | 365.3 (58.5) | 0.39* |

\*t test
\*\*Fisher's exact test
<sup>§</sup> Number of hours spent training for endurance events per week on average for the last six months

Table 5 compares participant demographics with and without ECG changes pre-race to post-race. Men tended to have more ECG finding change from pre-race to post-race than women, but this difference was not statistically significant (FET, *p* = 0.07). Years of training and past participation did not seem to affect ECG pre-to post-race. Increased number of hours of training per week seemed to favor change in ECG from pre to post-race. 34 (34.3%) of participants had sinus bradycardia pre-race. Of those, the bradycardia resolved in 24 of the 26 who presented for post-race ECGs. Eleven of the participants with sinus bradycardia pre-race did not have any other Seattle criteria present. However, other criteria were present post-race in 5 of those participants (sinus arrhythmia (1), incomplete RBBB (2), benign early repolarization (1) and isolated left ventricular hypertrophy (1)). Resolution of sinus bradycardia explained the conversion to “normal” ECGs in only 3 participants. Other common findings were increased QRS voltage significant for left ventricular hypertrophy (LVH) (24 (24.2 %) pre-race, 10 (14.1%) post-race), early repolarization (21 (21.2%) pre-race, 19 (26.8%) post-race) and incomplete right bundle branch block (RBBB) (8 (8.1 %) pre-race, 11 (15.5%) post-race).

## DISCUSSION

With increasing popularity of endurance events in the general population, more athletes may present to medical tents and emergency rooms (ED) with conditions such as chest pain, shortness of breath and syncope for which an ECG is a standard investigation. Physicians interpreting the ECG must determine if abnormalities are representative of athletic conditioning or a manifestation of an acute medical presentation. This may lead to either over or under investigation of the athlete. Minns et al [15] described two runners who presented to the ED with ECGs with a similar ischemic pattern consistent with acute myocardial infarction. Both underwent cardiac catheterization, one had a significant stenosis requiring stenting, the other had normal coronary arteries. This underscores the importance of careful history, physical exam, diagnostic testing including ECG and prudent use of invasive testing.

### Main findings

This observational study is one of the first looking at the acute effect of endurance activity on the ECG findings in a healthy asymptomatic athlete population. In a cohort of 99 athletes, 71.7% of ECGs pre-race were normal (≤ 1 Seattle Criteria C1-C10), including 36 (36.4%) with no Seattle criteria (C1-C26) present. Minns et al [2] reported similar findings: 27 of 87 (31%) athletes in their cohort had normal ECGs with no Seattle criteria present. The rest of the ECGs in their athlete population were considered clinically insignificant. We found 29 (29.2%) with the presence of more than 1 low-risk feature, most commonly sinus bradycardia (n=32), benign early repolarization (n=21), left ventricular hypertrophy (n=25) or both (n=9). Unlike the cohort reported by Minns et al.,[2] 5 (6.9%) athletes in our population had ECGs considered high-risk before the event. All were asymptomatic and had normal cardiac evaluations. Pelliccia et al.[20] examined a database of 12,550 trained athletes, and identified 81 with marked repolarization abnormalities and no apparent cardiac disease. Of the 81 athletes with abnormal ECGs, 5 (6%) ultimately proved to have cardiomyopathies. In comparison, 229 matched control athletes with normal ECGs from the same database did not develop cardiac disease or significant adverse events. They suggested abnormal ECGs in young and apparently healthy athletes may represent the initial expression of underlying cardiomyopathies and these athletes merit continued clinical surveillance. We did not observe any athletes with marked repolarization abnormalities in our cohort.

### Abnormal ECGs

We observed 28 (28.3%) of subjects with pre-event ECG findings considered abnormal as per the standards at the time.[6] With the recent consensus statement published in 2017 [21], the two subjects with isolated left axis deviation would not have been considered high risk and would not have required further investigation. No subjects developed concerning symptoms during training or racing consistent with prior studies suggesting a low predictive value of ECGs for cardiac abnormalities in athletes.[3] Interestingly, after exercise, more athletes had what is considered a “normal” ECG (83,3 %). This suggests an effect of exercise on the post-event ECGs. This “normalization” of ECGs during exercise was also observed by Minn et al.[2]

In our cohort, 17 subjects out of 72 with post-event ECGs had resolution of abnormal findings acutely after the endurance event. Post-event development of incomplete RBBBB was the most represented ECG change in this study. Development of long QT in 3 subjects was concerning and necessitated referral to cardiologists for further work-up. In these athletes, the corrected QT (QTc) was less than 500ms. Corrado et al [22] reported a 0.7% prevalence of long QT during their late screening period. Basavarajaiah et al. [23] investigated 2000 elite British athletes in 15 different sports for the prevalence and significance of an isolated long QT interval. They indicated the significance of an isolated prolonged QTc interval of <500 ms in athletes remains unknown but represents a low probability of long QT syndrome or a benign group in whom close monitoring rather than disqualification may be more appropriate in the absence a genetic diagnosis. Their research did not include the development of a long QT with exercise in athletes with previously normal QT intervals. More research in this area is indicated.

Our findings suggest participation in a long-distance aerobic event can acutely alter the ECG of asymptomatic athletes. With the growing popularity of endurance events, physicians may see an increase in athletes presenting to the Emergency room with symptoms warranting an ECG. It will be necessary for these physicians to interpret the findings taking into consideration the baseline ECG of the athlete and changes that may occur acutely related to event participation.

### Strengths and limitations

The strengths of our study are the use of a standardized approach to electrocardiograms acquisition and interpretation, using the AHA standards of acquisition. We were able to confirm with the on-site medical team that no participants presented for medical assessment.

Three reviewers blindly analyzed the ECG using the Seattle criteria, limiting bias. Our cohort is representative of high-volume athletic events as the participants were of different ages, sex and athletic performance levels.

However, our study was limited by the size of our cohort. Due to ethical and logistical limitations, we could not perform ECGs in symptomatic athletes: we would have needed to perform pre-race ECGs on a very large number of athletes to be able to contrast them to the few athletes who had cardiovascular related complaints at the finish line and this could have interfered with the provision of medical care.

### Application in the clinical setting

Emergency providers involved in the care of athletes during high-volume athletic events are challenged to interpret an athlete’s ECG and accurately differentiate findings suggestive of a potentially serious cardiovascular disorder from benign physiological adaptations occurring as the result of regular, endurance training.[21] ECGs are often performed on-site for participants who seek medical assistance for cardiovascular complaints such as syncope, dizziness, chest pain or palpitations, all of those complaints having multiple possible causes to explain them, cardiomyopathies, cardiac ischemia or arrhythmias being responsible for a small but relevant proportion.[22] Providers should be aware that high-volume exercise seems to affect ECGs and careful interpretation of ECGs obtained during athletic events is advised.

To further investigate the significance of ECG abnormalities in athletes, correlation to echocardiograms performed pre and post-race in this population might be of value. Also, obtaining ECGs in a symptomatic cohort of athletes with comparison to the baseline ECG would help interpret the significance of the findings. A larger scale study would confirm replication of these results.

## CONCLUSION

Long-distance endurance exercise might acutely affect the ECGs findings in asymptomatic athletes. We identified a high-prevalence of abnormalities in our cohort before exercise. Identifying physiologic electrocardiographic changes in athletes after completion of a high-intensity endurance event could help define new clinical guidelines for evaluation of cardiac complaints in athletes, since a significant proportion of them seem to have abnormalities on their baseline ECGs. Physicians providing care to long-distance athletes should interpret ECGs in this population prudently.

## Data Availability

Deanonymized datas available upon request.

## ACKNOWLEDGEMENTS

Marc Gosselin for permitting us to perform our study and Dave Boisvert, Pascale Caquez, Brendan Alexander and the new paramedics graduated from Ahuntsic College, for their help in recruitment and data collection. We would like to thank Sarah-Maude Martin and Christopher Bonneau Mercier, students in kinesiology from UQAM who identified athletes at the end of the race.

## SUPPLEMENTARY MATERIAL

Figure 1 – Flow Chart

Supplementary Material 1 – Demographic Questionnaire

Supplementary Material 2 – Management of ECG abnormalities pre-event and post-event

Supplementary Material 3 – Details of concerning ECG findings identified on site.

